# Blood biomarkers confirm subjective cognitive decline (SCD) as a distinct molecular and clinical stage within the NIA-AA framework of Alzheimer’s disease

**DOI:** 10.1101/2024.07.10.24310205

**Authors:** David Mengel, Ester Soter, Julia Maren Ott, Madeleine Wacker, Alejandra Leyva, Oliver Peters, Julian Hellmann-Regen, Luisa-Sophie Schneider, Xiao Wang, Josef Priller, Eike Spruth, Slawek Altenstein, Anja Schneider, Klaus Fliessbach, Jens Wiltfang, Niels Hansen, Ayda Rostamzadeh, Emra Düzel, Wenzel Glanz, Enise I. Incesoy, Katharina Buerger, Daniel Janowitz, Michael Ewers, Robert Perneczky, Boris Rauchmann, Stefan Teipel, Ingo Kilimann, Christoph Laske, Sebastian Sodenkamp, Annika Spottke, Johanna Brustkern, Frederic Brosseron, Michael Wagner, Melina Stark, Luca Kleineidam, Kai Shao, Falk Lüsebrink, Renat Yakupov, Matthias Schmid, Stefan Hetzer, Peter Dechent, Klaus Scheffler, David Berron, Frank Jessen, Matthis Synofzik, the DELCODE study group

**Affiliations:** Division Translational Genomics of Neurodegenerative Diseases, Hertie-Institute for Clinical Brain Research and Center of Neurology, University of Tübingen, Tübingen, Germany; German Center for Neurodegenerative Diseases (DZNE), University of Tübingen, Tübingen, Germany; German Center for Neurodegenerative Diseases (DZNE), Berlin, Germany; Charité – Universitätsmedizin Berlin, corporate member of Freie Universität Berlin and Humboldt-Universität zu Berlin,Institute of Psychiatry and Psychotherapy; Charité-Universitätsmedizin Berlin, Department of Psychiatry and Neurosciences, corporate member of Freie Universität Berlin and Humboldt-Universität zu Berlin, Berlin, Germany; German Center for Mental Health (DZPG), partner site Berlin; Charité – Universitätsmedizin Berlin, Experimental and Clinical Research Center (ECRC), Berlin, Germany; Department of Psychiatry and Psychotherapy, Charité, Berlin, Germany; School of Medicine, Technical University of Munich; Department of Psychiatry and Psychotherapy, Munich, Germany; University of Edinburgh and UK DRI, Edinburgh, UK; German Center for Neurodegenerative Diseases (DZNE), Bonn, Germany; Department for Cognitive Disorders and Old Age Psychiatry, University Hospital Bonn, Bonn, Germany; German Center for Neurodegenerative Diseases (DZNE), Göttingen, Germany; Department of Psychiatry and Psychotherapy, University Medical Center Göttingen, University of Göttingen, Göttingen, Germany; Neurosciences and Signaling Group, Institute of Biomedicine (iBiMED), Department of Medical Sciences, University of Aveiro, Aveiro, Portugal; Dept. of Psychiatry, Faculty of Medicine and University Hospital Cologne, University of Cologne, Cologne, Germany; German Center for Neurodegenerative Diseases (DZNE), Bonn/Köln, Germany; Excellence Cluster on Cellular Stress Responses in Aging-Associated Diseases (CECAD), University of Cologne, Cologne, Germany; German Center for Neurodegenerative Diseases (DZNE), Magdeburg, Germany; Institute of Cognitive Neurology and Dementia Research (IKND), Otto-von-Guericke University, Magdeburg, Germany; Department for Psychiatry and Psychotherapy, University Clinic Magdeburg, Magdeburg, Germany; German Center for Neurodegenerative Diseases (DZNE), Munich, Germany; Institute for Stroke and Dementia Research (ISD), University Hospital, LMU, Munich, Germany; Department of Psychiatry and Psychotherapy, University Hospital, LMU, Munich, Germany; Munich Cluster for Systems Neurology (SyNergy), Munich, Germany; Ageing Epidemiology Research Unit (AGE), School of Public Health, Imperial College London, London, UK; Sheffield Institute for Translational Neuroscience (SITraN), University of Sheffield, Sheffield, UK; Department of Neuroradiology, University Hospital LMU, Munich, Germany; German Center for Neurodegenerative Diseases (DZNE), Rostock, Germany; Department of Psychosomatic Medicine, Rostock University Medical Center, Rostock, Germany; Section for Dementia Research, Hertie Institute for Clinical Brain Research and Department of Psychiatry and Psychotherapy, University of Tübingen, Tübingen, Germany; Department of Psychiatry and Psychotherapy, University of Tübingen, Tübingen, Germany; Department of Neurology, University of Bonn, Bonn, Germany; Department of Old Age Psychiatry and Cognitive Disorders, University Hospital Bonn, Bonn, Germany; Department of Neurology, XuanWu Hospital of Capital Medical University, Beijing, China; Institute for Medical Biometry, University Hospital Bonn, Bonn, Germany; Berlin Center for Advanced Neuroimaging, Charité – Universitätsmedizin Berlin, Berlin, Germany; MR-Research in Neurosciences, Department of Cognitive Neurology, Georg-August-University Göttingen, Göttingen, Germany; Department for Biomedical Magnetic Resonance, University of Tübingen, Tübingen, Germany; Clinical Memory Research Unit, Department of Clinical Sciences Malmö, Lund University, Lund, Sweden

**Keywords:** blood biomarkers, plasma, phosphorylated tau, tau, neurofilament light chain, amyloid beta, magnetic resonance imaging, cognitive decline, longitudinal, early Alzheimer’s disease, preclinical Alzheimer’s disease

## Abstract

**Introduction:** Subjective cognitive decline (SCD) is proposed to indicate transitional stage-2 in the AD continuum, yet longitudinal fluid biomarker data for this stage is scarce. We investigated if blood-based biomarkers in amyloid-positive individuals with SCD (A+SCD) support stage-2 as distinct from AD stages-1 and -3 and identify those at high risk for progression.

**Methods:** We analyzed plasma phospho-tau-181 (p181) and neurofilament-light-chain (NfL) in a prospective multicenter study of 460 participants across the AD continuum, assessing their association with cognition, hippocampal atrophy, and clinical progression.

**Results:** Baseline plasma p181 was elevated and increased faster in A+SCD compared to amyloid-positive cognitively unimpaired (A+CU) individuals (stage-1). NfL rose across A+CU, A+SCD, and A+MCI (stage-3). In A+SCD, higher p181 predicted cognitive decline and transition to MCI.

**Discussion:** Plasma p181 provides biomarker evidence for A+SCD as a distinct pre-dementia AD stage and helps identify individuals at risk for cognitive decline early in the AD continuum.

**Research in Context:** *Systematic Review:* Research on subjective cognitive decline (SCD) and its association with Alzheimer’s disease (AD), as well as investigations into stage-2 of the AD continuum, is quickly expanding, but fluid biomarker evidence is scarce. We conducted a comprehensive review across PubMed, recent meeting abstracts, and oral presentations, focusing on cross-sectional and longitudinal case-control studies, cohort studies, and meta-analyses.

*Interpretation:* Our plasma phospho-181 tau (p181) findings provide molecular fluid biomarker evidence for A+SCD as a pre-dementia AD stage (stage-2) distinct from A+CU (stage-1). Plasma p181 assessment aids in identifying individuals at risk of future disease progression early in the AD continuum.

*Future directions:* The here proposed concept of SCD as an indicator of stage 2 of the Alzheimer’s disease continuum - supported and stratified by easily accessible blood-based biomarkers - warrants further validation in memory clinics. It could facilitate earlier- and thus even higher-effect - treatments in the pre-dementia stages of AD.

*Highlights:*

- A+SCD exhibits a distinct trajectory of plasma p181 compared to A+CU
- Higher plasma p181 levels in A+SCD predict PACC5 decline and transition to MCI
- Plasma p181 serves as a biomarker that delineates the A+SCD stage from A+CU Plasma p181 levels stratify SCD patients, facilitating early interventions

## 1. Introduction

The pathophysiological processes of Alzheimer’s disease (AD) including amyloid and tau deposition unfold many years prior to the appearance of initial symptoms and subsequent progression to dementia [1]. This prolonged pre-dementia period presents a crucial window for potential interventions to be introduced, aiming to alleviate or defer the onset of cognitive decline [2–5]. The National Institute on Aging and Alzheimer’s Association (NIA-AA) working group’s updated research criteria delineated 6 progressive clinical stages evident in individuals along the AD continuum, which is identified by the presence of amyloid pathology with or without tau pathology [6]. Within the pre-dementia stages, this staging system includes a transitional stage 2 positioned between the fully asymptomatic stage 1 and stage 3 defined by mild cognitive impairment (MCI). A symptom that has been associated with stage 2 is subjective cognitive decline (SCD) [6, 7]. SCD is defined as self-experienced decline in cognitive functioning, while - in contrast to MCI - the performance on diagnostic neuropsychological tests is normal. In elderly individuals, SCD is associated with an increased risk of future cognitive decline [8], and may occur more than 15 years before dementia onset [9, 10]. Meta-analyses reported a conversion rate from SCD to MCI and dementia in 27% and 6-14 %, respectively [11, 12]. Recent clinical research findings indicate that individuals experiencing subjective cognitive decline in the presence of amyloid pathology (A+SCD) show reduced cognitive and functional performance and face a significantly increased risk of progressing to MCI and dementia when compared to SCD individuals without AD pathology [13, 14], providing evidence that A+SCD might present a distinct pre-dementia AD disease stratum. However, there is a scarcity of comprehensive characterizations of molecular changes in biofluids (in particular blood) of individuals with subjective cognitive decline in incipient AD (A+SCD), including their longitudinal trajectories; in particular in their long-term longitudinal association with cognitive decline and brain alterations, and when specifically compared to both cognitively unimpaired individuals without cognitive decline (A+CU) and those with cognitive impairment in the presence of amyloid pathology (A+MCI) [8]. Investigations into molecular characteristics of SCD are essential to validate the concept of A+SCD as an indeed distinct disease stratum, serving as the transitional stage 2 between fully asymptomatic individuals (stage 1) and MCI (stage 3). This might allow to further strengthen the conceptual relevance of SCD in AD, and also to deliver specific molecular biomarker signatures - even in blood-for both prediction of clinical progression and stratification of participants for early intervention. This is of immediate importance because an increasing number of disease-modifying AD therapies is under approval or in development [4, 15, 16].

Blood-based biomarkers are ideally suited for tracking molecular changes longitudinally and predicting clinical outcomes in pre-dementia AD stages including SCD, given their extensive applicability, minimal invasiveness, and the ease with which they can be repeatedly assessed over time [17], in particular when compared to PET or CSF. Blood biomarker studies have demonstrated that - amongst other plasma tau markers - tau phosphorylated at threonine-181 (p181) specifically captures AD brain pathology, and can differentiate AD from other neurodegenerative diseases [2, 18]. Furthermore, plasma p181 has been shown to predict cognitive decline and progression from MCI to dementia [19]. Neurofilament light chain (NfL), a non-disease specific marker for the intensity of ongoing axonal damage, has also demonstrated potential as a blood-based biomarker in neurodegenerative diseases including AD [20, 21]. Elevated plasma NfL levels have been associated with cognitive decline and an increased risk for progression from MCI to dementia [22, 23].

In this study, we aimed to explore the role of plasma p181 tau and neurofilament light (NfL) levels as biomarkers for T (tau pathology) and N (neurodegeneration) within the ATN framework of AD pathology [6] in SCD. Specifically, we investigated whether these biomarkers and their changes over time could serve as molecular evidence for SCD as a distinct stage in the progression of AD. Additionally, we examined whether these biomarkers could help identify individuals at higher risk for future clinical deterioration.. To this end, we assessed plasma p181 tau and NfL levels and their longitudinal trajectories in their association with hippocampal atrophy, prospective cognitive decline, and AD stage transition to MCI and dementia in a large prospective multi-center cohort of individuals with SCD. The observed changes were specifically contrasted to the alterations in both CU and MCI individuals, thus allowing to validate SCD as a distinct clinical and molecular stage in the pre-dementia AD continuum.

## 2. Material and Methods

### 2.1 Participants

We analyzed data from 457 participants of the Longitudinal Cognitive Impairment and Dementia Study (DELCODE) of the German Center for Neurodegenerative Diseases (DZNE), of whom CSF data was available. DELCODE is an observational longitudinal memory clinic-based multicenter study carried out by DZNE associated university memory clinics in Germany. The ethical committees of all participating centers approved the protocol. All participants provided informed consent prior to study participation. A complete description of all inclusion and exclusion criteria has been published elsewhere [24]. In brief, all participants were 60 years or older and enrolled between 2014 and 2018. For this study, we included all patients from which plasma samples as well as baseline CSF was available. All participants classified as SCD (n=210) presented to memory clinics through referral or self-referral with complaints of cognitive decline, and fulfilled the SCD research criteria (i) self-experienced decline in cognitive functioning, compared with a previously normal cognitive status, which is unrelated to an acute event; and (ii) normal performance on standardized tests used to classify MCI, corrected for age, sex, and education. Unimpaired cognition in objective testing was defined as a test performance better than −1.5 SD on all subtests of the Consortium to Establish a Registry for Alzheimer’s Disease (CERAD) neuropsychological test battery. The non-SCD cognitively unimpaired group (CU, n=89) was recruited by advertisement, explicitly addressing individuals who felt healthy and without any relevant cognitive problems. Unimpaired cognition of CU individuals was verified according to the same criteria as above described for the SCD group. In addition, participants with amnestic MCI (n= 110) and mild dementia of Alzheimer type (DAT) (n= 48, MMSE ≥ 18 points) were recruited according to current research criteria for MCI and DAT (NIA-AA) [25, 26].

### 2.2 Cognitive testing

At baseline and annual follow-ups a comprehensive cognitive test battery was applied by trained neuropsychologists at all sites [24, 27]. We calculated the Preclinical Alzheimer Cognitive Composite (PACC5), which has been extensively validated to detect subtle cognitive changes in individuals who are in the pre-dementia stage of Alzheimer’s disease [28, 29]. PACC5 was calculated as the average z-standardized performance in memory (FCSRT Free Recall and Total Recall), verbal episodic memory (Wechsler Memory Scale – Forth Edition (WMS-IV) Logical Memory Story B delayed recall), global cognition (MMSE),, attention and processing (Symbol-Digit-Modalities Test), and verbal fluency (the sum of two category fluency tasks). Baseline mean and SD values of the CU group were used to derive the subtest z-scores. In CU and SCD, incident MCI was diagnosed by clinical consensus in individuals with evidence of longitudinal cognitive decline [30]. In MCI patients, incident dementia was diagnosed by the study physician according to established criteria.

#### Magnetic resonance imaging

Magnetic resonance imaging (MRI), including up to 4 years longitudinal imaging, was carried out at nine imaging sites on Siemens 3T-MR-Scanners according the DZNE imaging protocol, and quality control process, as described previously [24]. Automatic hippocampal subfield segmentation was performed on high-resolution T2-weighted images, from which whole hippocampal volumes were derived using the Freesurfer image analysis suite [14], which is freely available for download online (http://surfer.nmr.mgh.harvard.edu/).

### 2.4 Fluid biomarkers

#### CSF biomarkers

CSF Aβ42, Aβ40, and total-tau were analyzed centrally in one lab using commercial V-Plex ELISAs (Mesoscale Diagnostics, Rockville, USA); and CSF p181 with the Innotest Phospho-Tau(181P) ELISA (Fujirebio Germany GmbH, Hannover, Germany). Independent reference samples were used to control assay performance. Cut-off values for both CSF Aβ42/Aβ40 ratio and tau were determined from the DELCODE data set using Gaussian mixture modelling using the R package flexmix, version 2.3-15 [14]. The following cut-offs were applied to indicate Alzheimer’s pathological changes: CSF Aβ42/40 ratio ≤ 0.08 and total-tau ≥ 510.9 pg/mL. The cut-off of the Aβ42/40 ratio was used to define amyloid positivity. We aimed to examine the trajectory of plasma biomarkers and their association with clinical measures in individuals across the AD pre-dementia continuum. Following the 2018 NIA-AA research framework, Aβ biomarker positivity indicates whether an individual falls in the AD continuum. Participants were thus grouped based on their CSF amyloid levels, including: participants with amyloid deposition, but without cognitive impairment and withoutsubjective cognitive decline (A+CU); participants with amyloid deposition and subjective cognitive decline (A+SCD); and participant with amyloid deposition and mild cognitive impairment (A+MCI). Clinical groups without evidence of Alzheimer’s pathology (A-CU, A-SCD, and A-MCI) were included for comparison (Tab. 1 and Fig. S1 A+B).

**Table 1:**
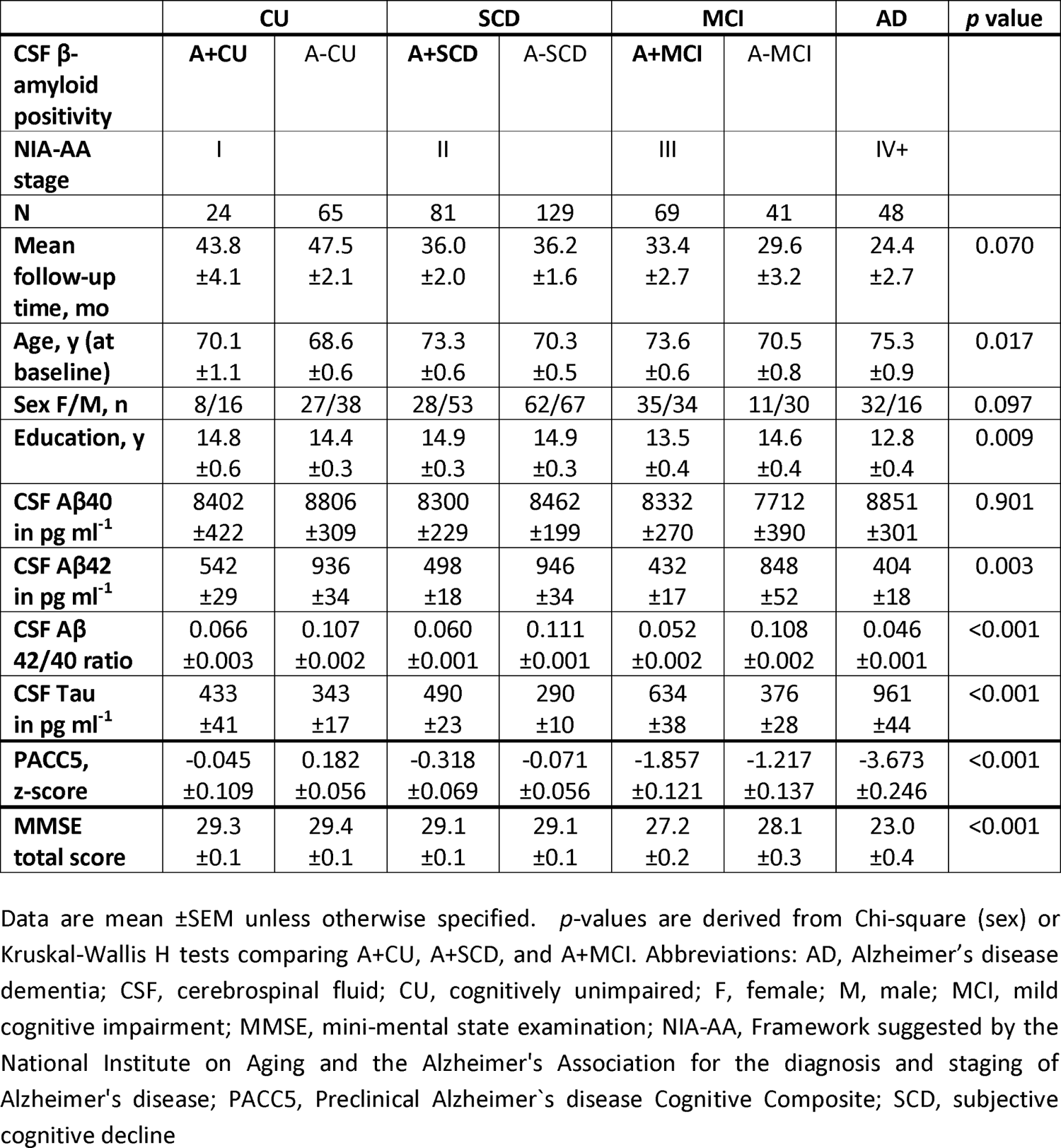
Demographics and clinical characteristics.

#### Plasma biomarkers

Plasma levels of p181 and NfL were quantified using Simoa assays. All assays were performed by the same operator, and conducted on Quanterix HD-1 and HD-X instruments (Quanterix, Billerica, MA). Simoa NF-light Advantage (Quanterix, Billerica, MA, USA) and pTau-181 advantage version 2 kits were used according to the manufacturer’s instructions. Samples were diluted 1:4 in sample buffer and analyzed in technical duplicates. The lower limit of quantitation (LLoQ) was defined as the lowest standard: (i) with a signal higher than the average signal for the blank plus 9 SDs, and (ii) allowing a percent recovery ≥100 ± 20 %. For p181 and NfL, the LLoQ between runs was 0.77 pg/mL and 1.80 pg/mL, respectively. Two internal control samples were assessed both at the start and end of each assay run to determine repeatability and inter-assay variability. The % repeatability for p181 and NfL was 5.6% (sample 1) and 7.6% (sample 2), and 9.2% (sample 1) and 4.6% (sample 2), respectively.

The inter-assay variance was 12.4% (sample 1) and 11.8% (sample 2), and 9.1% (sample 1) and 6.5% (sample 2), respectively. Samples were excluded from further analysis (2.4% for p181, and 3.8% for NfL), if the %CV was > 20% between two technical replicates or if measurements were available from of only one technical replicate.

### 2.5 Statistical analysis

Statistical analyses were carried out using GraphPad Prism, version 10.1.2 (LaJolla, CA, USA) and Stata, version 17.0 (College Station, TX, USA). Normal distribution was assessed by visual inspection of histograms and Quantil-Quantil-plots. Differences of biomarker levels between groups were assessed using Welch-ANOVA followed by Dunnetts post hoc test (for normally distributed p181 data) or Kruskal Wallis H test followed by Dunn’s post hoc test (for non-normally distributed NfL data). Linear mixed effect (LME) models were fitted to analyze plasma p181 and NfL trajectories over time in CU, SCD, and MCI individuals stratified by amyloid positivity. LME models were adjusted for age and gender, and included an interaction between time and diagnostic group, as well as random intercepts and slopes nested within subject. To study associations of amyloid-positivity or baseline plasma levels (categorized by 3-quantiles) with longitudinal PACC5 and hippocampal volume (average of left and right sight), LME models were fitted including an interaction factor between the categorical variable of interest and time. Models were adjusted for age and sex, and included random intercepts and slopes nested within subject. For cognition, we also included years of education as covariate. Total brain volume was incorporated as a covariate for hippocampal volume analysis. Linear additivity was assessed by visual inspection of the residuals vs. fitted plot. *p*-values were adjusted for multiple comparisons through false discovery rate (FDR) using the Benjamini-Hochberg correction [31, 32] (if more than 2 comparisons were examined for an interferential hypothesis), and were considered significant at p≤0.05, two-tailed. To assess associations between amyloid positivity and plasma levels of p181 and NfL (categorized by 2-quantiles) and risk of incident MCI or dementia, we used Kaplan-Meier survival analysis and cox proportional hazards regression. The proportionality of hazards was assessed using the Schoenfeld residuals.

## 3. Results

### A+SCD individuals exhibit a distinct plasma p181 trajectory and increased plasma NfL levels

Cross-sectional plasma p181 levels were elevated in A+SCD compared to A+CU (1.7±0.1 vs 1.1±0.1 pg/mL; p=0.009) and to A-SCD (1.1±0.0 pg/mL; p<0.001) (Fig. 1A), while there was no difference of A+SCD compared to A+MCI (1.9±0.1 pg/mL; p=0.453) (for comparison to CSF p181 levels, see Supplement 1).Plasma NfL levels were increased in A+SCD compared to A-SCD (17.7±1.3 vs 14.3±0.6 pg/mL; p=0.022). Compared to A+CU, plasma NfL levels in A+SCD showed a numeric (Fig. 1) but statistically non-significant elevation (17.7±1.3 vs 13.7±1.2 pg/mL; p=0.276), and NfL levels were then further elevated in A+MCI (22.0±1.6 pg/mL) compared to A+SCD (p=0.008) (for stepwise increase pattern across the three pre-dementia AD stages, see Fig. 1).

**Figure 1.**
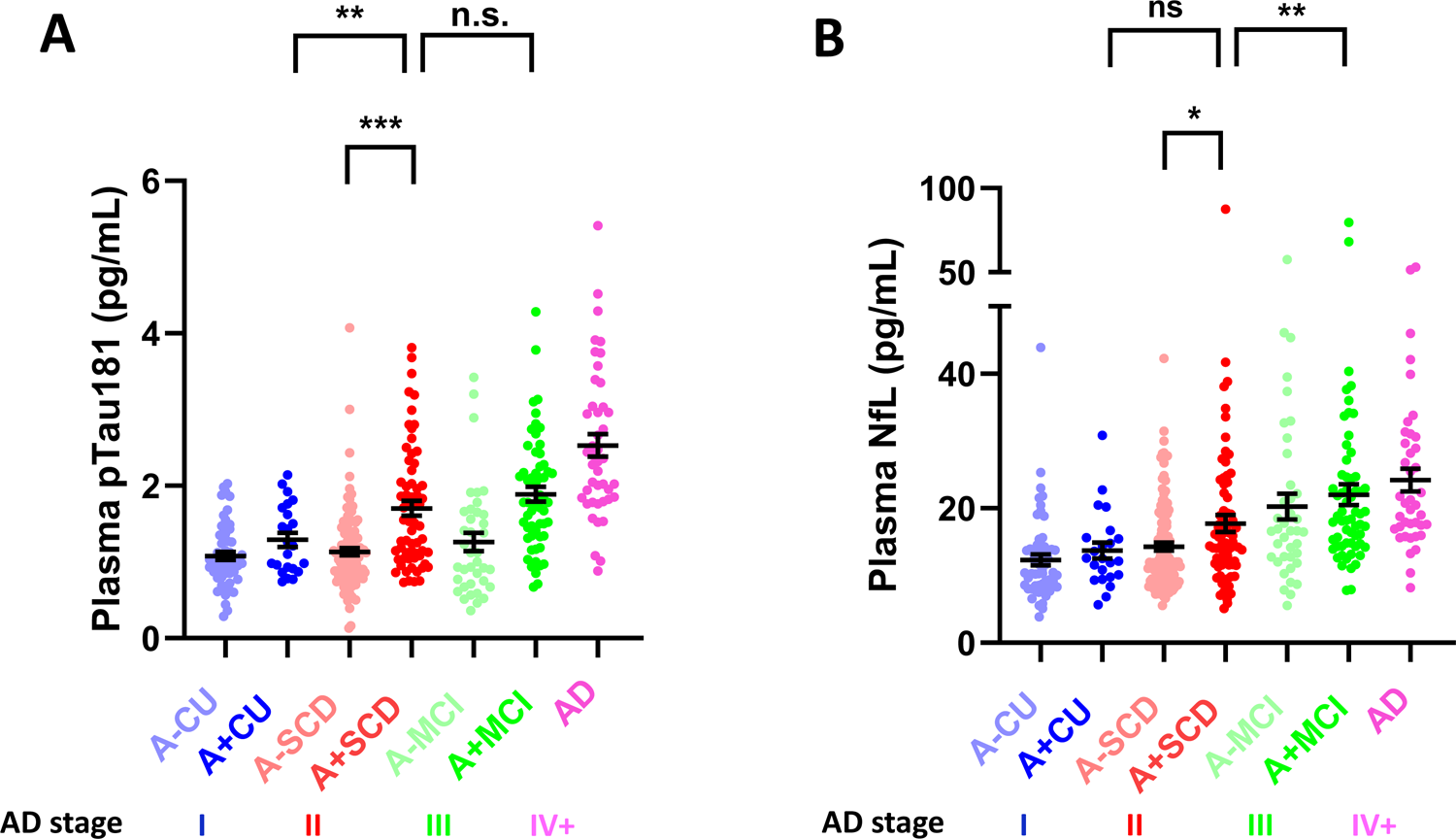
Plasma p181 and NfL are elevated in A+SCD. CU (blue), SCD (red), and MCI (green) individuals were stratified by CSF-amyloid positivity (A+ vs A-), and their baseline plasma analyzed using Simoa assays to detect **(A)** p181 **(B)** NfL. Plasma from AD patients (purple) was included as a reference for both biomarkers. The respective stage in the AD continuum, operationalized using clinical assessment combined with CSF amyloid-positivity, is indicated below the graph (I: A+CU, II: A+SCD, III: A+MCI, IV+: AD). In both graphs, each point represents an individual subject, and means ± SEM are indicated. Differences between groups were assessed using Welch-ANOVA followed by Dunnetts post hoc test (for p181 data) or Kruskal Wallis H test followed by Dunn’s post hoc test (for NfL data)., \**p*<0.05, \*\**p*<0.01, \*\*\**p*<.001, n.s., non-significant. **(A)** Plasma p181 levels are elevated in A+ SCD compared to A+CU and A-SCD, while levels do not differ between A+SCD and A+MCI. **(B)** Plasma NfL levels show a trend towards increased levels in A+SCD compared to A+CU, and are significantly icreased in A+SCD compared to A-SCD. Plasma NfL levels are then further elevated in A+MCI vs A+SCD.

Analysis of longitudinal change demonstrates that plasma p181 levels increase at a faster rate over time in A+SCD (0.15 pg/mL/year) compared to both A+CU (0.05 pg/mL/year) (slope A+SCD vs A+CU, p=0.044) and A-SCD (0.06 pg/mL/year) (slope A+SCD vs A-SCD, p=0.016) individuals (Fig. 2), indicating that A+SCD individuals have a distinct trajectory of molecular ptau pathology, which is different from both A+CU and A-SCD. This accelerated rate of change, starting off in the SCD stage, is then sustained in A+MCI (slope A+SCD vs A+MCI, p=0.308).

**Figure 2.**
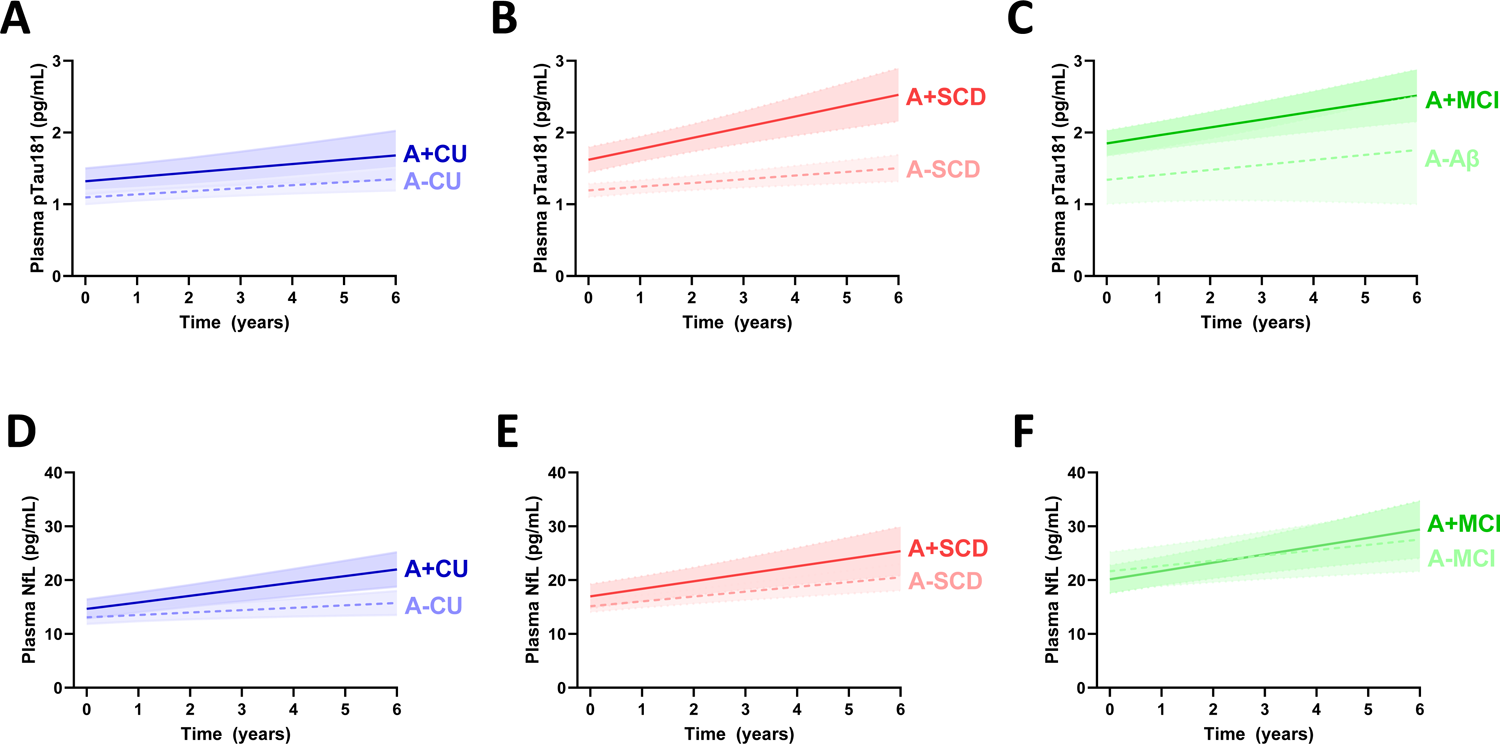
Plasma p181 levels increase at a higher rate over time in A+SCD and A+MCI compared to A+CU, while plasma NfL levels rise at a similar rate in A+CU, A+SCD, and A+MCI. Plasma p181 and NfL was measured in plasma samples collected at ∼1 year intervals from **(A + D)** A+CU and A-CU, **(B + E)** A+SCD and A-SCD, and **(C + F)** A+MCI and A-MCI. **(A + B)** Plasma p181 levels in A+ SCD, compared to each A+CU and A-SCD increase more steeply over time. **(C)** The rate of change is then kept in MCI. **(D – F)** Changes of plasma NfL levels over time are similar in A+CU, A+SCD, and A+MCI, which leads to elevated NfL in A+SCD and A+MCI compared to A+CU individuals. Estimated trajectories for CU (blue), SCD (red), and MCI (green) are drawn using mixed-effects modelling with an interaction term for time and amyloid-positivity, and adjusted for age at baseline, and sex. Shaded areas indicate 95% confidence intervals.

Longitudinal changes in plasma NfL levels were similar in A+CU (1.22 pg/mL/year), A+SCD (1.40 pg/mL/year), and A+MCI (1.54 pg/mL/year) individuals (slope A+SCD vs A+CU, p=0.790; slope A+SCD vs A+MCI, p=0.790), indicating a similar axonal turnover rate across the pre-dementia AD continuum (slope A+CU vs A+SCD vs A+MCI, p=0.765).

### Baseline plasma p181 levels predict future cognitive decline in A+SCD

The rate of cognitive decline over time, as assessed through longitudinally measured PACC5, was increased already in individuals with SCD (slope SCD vs CU, p=0.001), with this cognitive trajectory mainly driven by the stronger cognitive decline in A+SCD (−0.06±0.03 units/year) than A-SCD individuals (+0.01±0.02 units/year) (slope A+SCD vs A-SCD, p=0.022, Fig. 3). In contrast to SCD, no cognitive deterioration could be observed in the CU group (+0.05±0.01 units/year), also not in the A+CU group (+0.04±0.02 units/year), as shown before in this cohort [14]. This cognitive decline, starting off in the SCD stage, was accelerated further in MCI (−0.25±0.04 units/year) compared to SCD (slope MCI vs SCD, p<0.001), with the greatest decline observed in A+MCI individuals (−0.30±0.06 units/year, slope A+MCI vs A+SCD, p=0.005).

**Figure 3.**
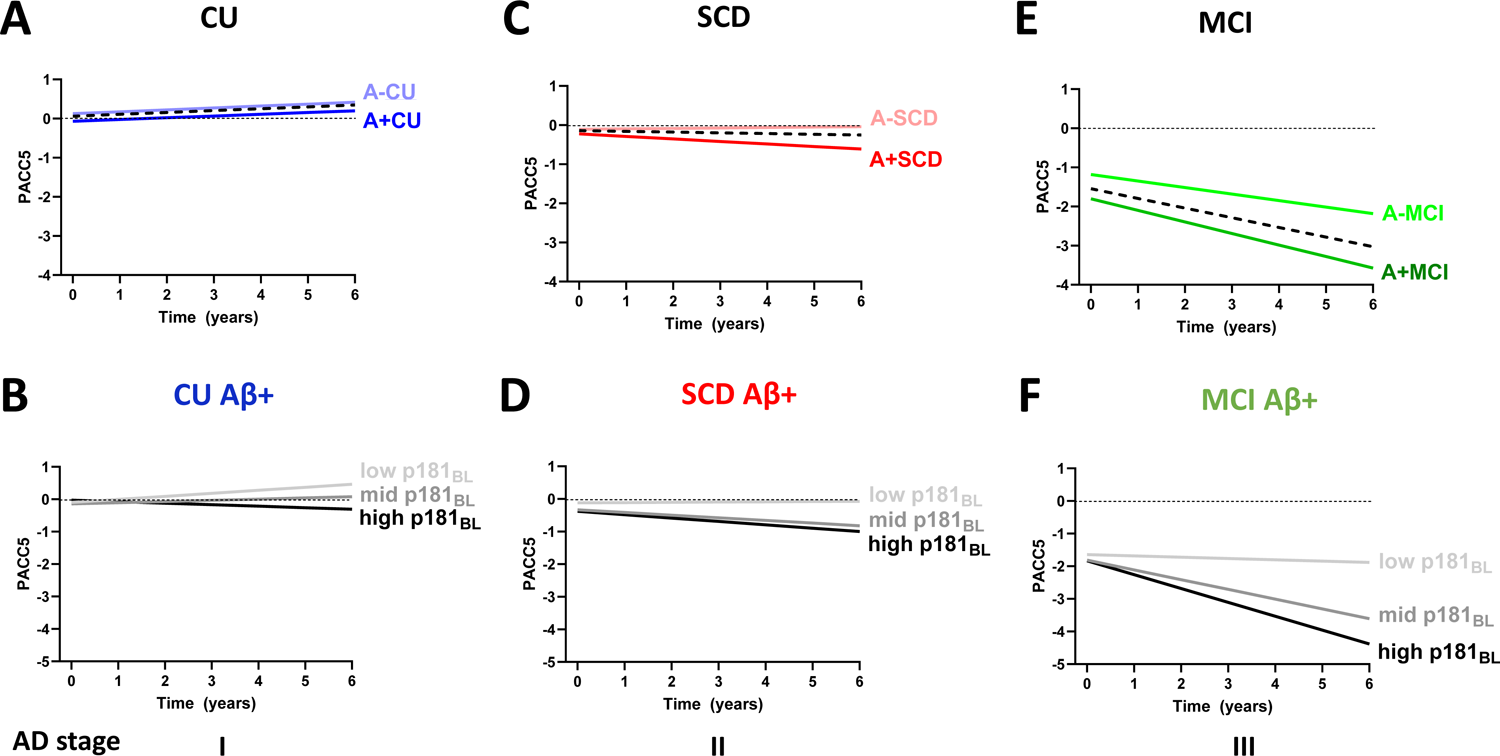
Longitudinal cognitive decline in A+SCD is predicted by higher plasma p181 levels at baseline. Longitudinal trajectories of the PACC5 score of **(A)** CU (blue), **(B)** SCD (red), and **(C)** MCI (green) individuals, stratified by CSF amyloid-positivity are displayed. Dashed black lines indicate the trajectories for the clinical groups (CU, SCD, MCI) irrespective of the CSF amyloid status. **(A – C)** The rate of cognitive deterioration over time, as measured by PACC5, is increased in SCD compared to CU, largely driven by the worse performance of A+SCD individuals. The observed cognitive trajectory in A+SCD is then accelerated further in A+MCI. In panel D – F, baseline (BL) plasma p181 levels of the A+CU, A+SCD, and A+MCI group, categorized into low (light grey), mid (medium grey), and high (dark grey) levels using 3-quantiles, are used to predict longitudinal performance on PACC5. **(D – E)** Future cognitive decline observed in the A+SCD group is associated with high p181 levels at baseline. This predictive association is not seen to the same extent in the A+CU group. Once this association is started in the A+SCD, it is then maintained in the A+MCI group. Trajectories were derived from linear mixed models with an interaction term for time and baseline p181 levels, and adjusted for age at baseline, sex, and years of education.

In the A+SCD group, higher plasma p181 levels at baseline were associated with cognitive decline (p=0.036). In contrast, this association was not observed to a similar extent in the A+CU group. A standardized annual PACC5 change of 0.10±0.03 was noted in individuals with A+SCD exhibiting the highest p181 levels, in contrast to an annual change of only 0.05±0.02 for the A+CU group with the highest p181 levels. Once starting off in the A+SCD group, the association between higher baseline p181 levels and subsequent cognitive deterioration is then maintained and further accelerated in the A+MCI group (Fig. 3). In contrast to p181 levels, baseline NfL levels were associated with cognitive decline in A+MCI individuals, but not yet in A+SCD individuals (Fig. S2).

We also examined hippocampal volume loss in the AD pre-dementia stages, including up to 4 years longitudinal volumetric MRI, in relation to blood biomarker levels. At baseline, hippocampal volumes were lower in SCD compared to CU, and further reduced in MCI (Fig. S3).. In A+MCI, but not yet in A+SCD, higher baseline ptau181 levels (Fig. S3), were numerically associated with lower hippocampal volume, while NfL levels (Fig. S4) did not show any trend of association. Further discussion on these findings is provided in the Supplement.

### Baseline levels of plasma p181 predict progression to MCI in A+SCD

A total of 202 SCD and 94 MCI individuals were evaluated for clinical progression to MCI and dementia at follow-up, respectively. Of those, 44 SCD (21.8%) and 37 MCI (39.4%) converted to MCI or dementia, respectively. A+SCD individuals had a higher risk of progression to MCI than A-SCD individuals (hazard ratio 1.9 ±0.6, p=0.039) (Fig. 4). In a mean follow-up time of 3.0±0.1 years, 29.5% of A+SCD converted MCI, compared to 16.9% in the A-SCD group. Mean time to conversion was 2.3±0.2 years for converters from SCD to the MCI stage. Similarly, A+MCI individuals had a higher risk of progression to AD than A-MCI individuals (hazard ratio 3.1±1.4, p=0.011) (Fig. 4).

**Figure 4.**
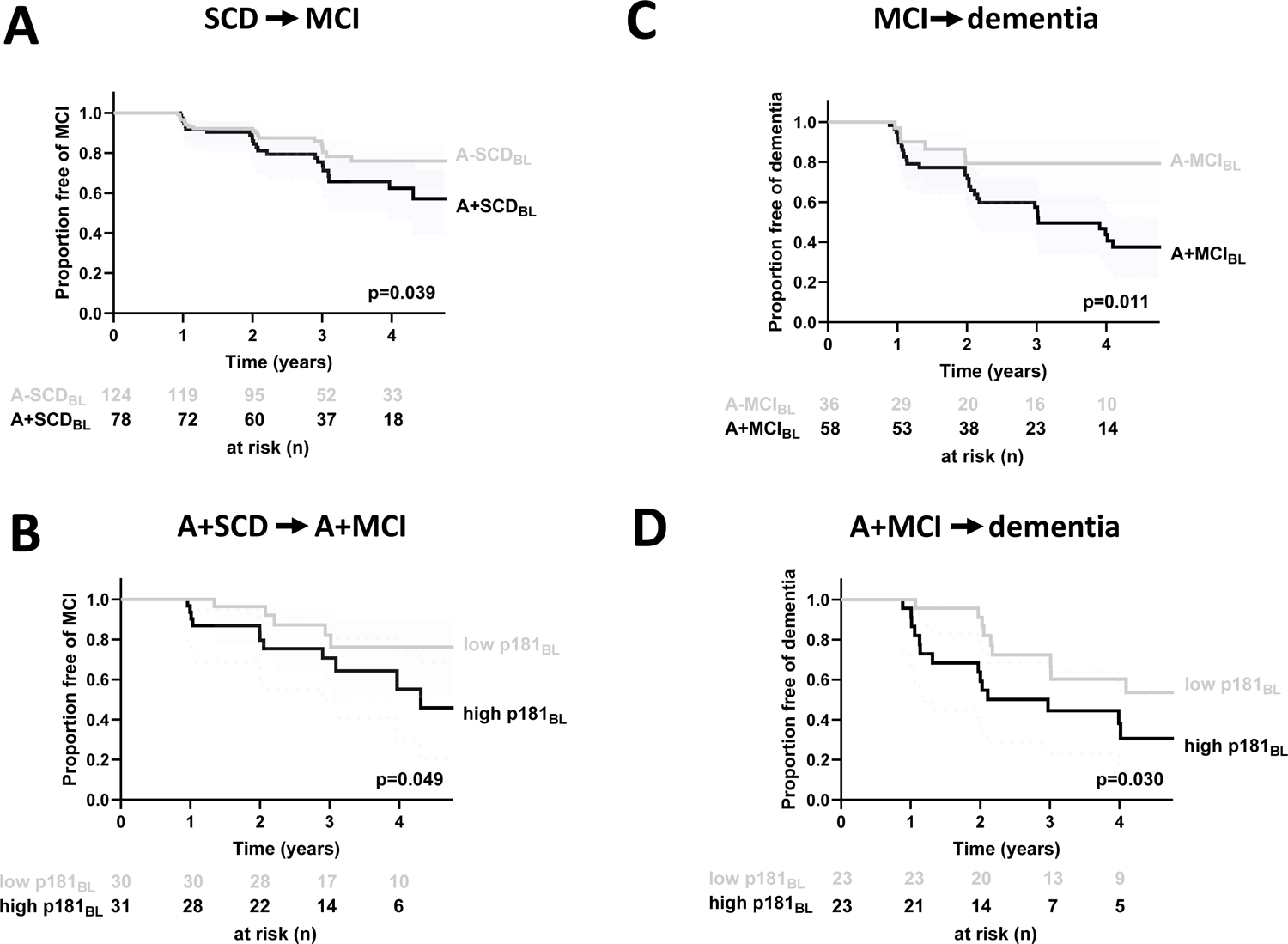
A+SCD compared to A-SCD individuals have a higher risk for conversion to the MCI stage, which is further increased in the presence of higher baseline levels of p181. Survival curves for progression from **(A)** SCD to MCI, and **(B)** MCI to dementia, stratified by CSF amyloid-positivity are indicated. **(A)** SCD and **(B)** MCI individuals have a higher risk of progression to MCI and AD, respectively, in the presence of amyloid deposition. In panel C and D, progression from **(C)** A+SCD to MCI, **(D)** and A+MCI to dementia among individuals with high (first and second quartile, in black) versus low (third and fourth quartile, in light grey) baseline plasma levels of p181 are shown. The x-axis shows the time of follow-up from diagnosis of SCD or MCI, while the y-axis is the fraction of patients free of MCI or dementia at given time-points. Values below each graph indicate the number of subjects free of MCI/dementia at 0, 1-, 2-, 3-, and 4-years follow-up from first detection of SCD/MCI. Higher baseline plasma p181 levels predict the conversion of **(C)** A+SCD to MCI, and **(D)** A+MCI to dementia.

Higher baseline p181 levels were significantly associated with future conversion from SCD to MCI (hazard ratio=2.9±1.5, p=0.049) in A+SCD. Similarly, higher baseline p181 levels were also significantly associated with future progression from A+MCI to dementia (hazard ratio=2.5±1.1,p=0.030). Baseline NfL levels did not significantly predict clinical AD stage transition (Fig. S5).

## 4. Discussion

This prognostic study investigated the potential of blood-based biomarkers, specifically plasma p181 and NfL, to reveal molecular changes in individuals with SCD. It would provide molecular blood biomarker support for SCD as a transitional stage 2 between the fully asymptomatic stage 1 and stage 3 according to the NIA-AA Research Framework; plus – at the same time-pave the path for a blood-based signature of the ATN AD scheme (with p181 representing T/tau, and NfL representing N/neurodegeneration), thereby also facilitation the NIA-AA’s 2023 proposal to include blood-based biomarker in this classification scheme [33]. Moreover, we also examined their capacity to predict both future cognitive decline and AD stage transition to MCI.

### Plasma p181 is elevated in A+SCD

Our results reveal an elevation in baseline plasma p181 levels in A+SCD in comparison to A+CU, and this elevation also increases at a higher rate over time compared to A+CU individuals. While individuals in both A+CU and A+SCD groups lack cognitive impairment and, consequently, appear indistinguishable in neuropsychological assessments, the biomarker trajectories of plasma p181 in A+SCD more closely resemble those observed in A+MCI than in A+CU. This suggests a pertinent molecular progression along the pre-dementia AD continuum in individuals with SCD (stage 2), to a similar extent as in A+MCI (stage 3), but distinct from those without subjective cognitive decline (A+CU, stage 1). Prior research has shown a rise in plasma phospho-tau including p181 along the clinical AD continuum [34], with elevated levels observed in AD dementia compared to both MCI and CU [19, 35]. It has been proposed that the release of extracellular soluble tau is related to early dysregulation in neuronal tau metabolism due to early Abeta pathology, and (in later stages) tau fibril formation [34]. However, when assessing plasma ptau levels in AD and MCI patients, the comparisons were largely made with cognitively unimpaired individuals or healthy controls. Furthermore, SCD patients were often times placed within either the CU or MCI groups, precluding to determine the role of biomarkers in this particular pre-dementia AD stratum [19, 20]. Notably, to our knowledge no study has so far explored longitudinal ptau biomarker trajectories in A+CU in direct comparison to A+SCD groups. This study addressed this gap and enabled us to demonstrate a unique molecular ptau trajectory in individuals with A+SCD. The observed elevation in p181 levels in A+SCD, before the onset of objective cognitive impairment, underscores the potential utility of this blood-based biomarker in identifying individuals at this very early stage of the pre-dementia AD continuum.

### Plasma NfL adds further support for SCD stage an intermediate AD stage between CU and MCI, with incipient axonal degeneration

Plasma NfL levels were tended to be numerically increased levels in A+SCD compared to A+CU, and were - in particular-significantly increased in A+SCD compared to A-SCD. Once starting off at the A+SCD stage, plasma NfL levels then increased further in the A+MCI stage. Within the SCD cohort, these findings suggest that axonal degeneration is already detectable and peripherally captured in A+SCD individuals. This molecular biomarker signature of early incipient neurodegeneration from the SCD stage onwards in the clinical AD continuum further supports A+SCD as an intermediate between fully asymptomatic A+CU (= AD stage 1) and A+MCI (= AD stage 3). In contrast, NfL biomarker analysis does not support A-SCD as a neurodegenerative state. This differs from the MCI stage, where individuals –irrespective of their amyloid status – show a molecular biomarker signature of axonal degeneration (i.e. increased NfL levels in both A+ and A-MCI, without statistical difference). This sequential elevation in NfL levels across the pre-dementia AD stages further supports the idea of a continuous biological process underlying the progression of AD along the proposed clinical continuum. This is further supported by the fact that longitudinal changes in plasma NfL levels were similar in A+CU (stage 1), A+SCD (stage 2) and A+MCI (stage 3), indicating a similar axonal turnover rate across the pre-dementia AD continuum.

In contrast to the distinctive plasma p181 trajectory observed in individuals with A+SCD, alterations in NfL levels at baseline and over time between A+CU and A+SCD are, however, more nuanced, merely reflecting a trend - and likely a lesser degree of neurodegeneration at these very early stages of pre-dementia AD compared to MCI.

### Plasma p181 predicts future cognitive decline in A+SCD

The rate of cognitive decline over time, as assessed by PACC5, was accelerated in individuals with SCD, primarily driven by the poorer performance in A+SCD. In contrast to SCD, the CU group exhibited no cognitive decline, regardless of amyloid positivity. This cognitive trajectory, starting off in A+SCD individuals, was then further accelerated in A+MCI. These findings, supporting earlier reports from the DELCODE cohort [14], indicate that A+SCD shows a distinct cognitive trajectory different from CU, including also in particular from A+CU. This degree of future cognitive decline in the A+SCD group could be predicted by plasma p181 levels at baseline. In contrast, this predictive association was not seen to the same extent in the A+CU group. Once this association is started in the A+SCD, it is then maintained in the A+MCI group. Taken together, this thus adds further support for A+SCD as an intermediate stage between A+CU and A+MCI.

This is the first study showing a specific association of plasma p181 with longitudinal cognition in A+SCD vs. A+CU individuals. In addition, our data also extend and specify previous studies that had indicated a relationship between baseline phosphorylated tau measures in CSF and future cognitive decline in SCD [13, 36]. The now identified predictive association of elevated plasma p181 levels in A+SCD for future decline in PACC5 holds important clinical significance. These findings indicate that plasma p181 levels may function as a predictive marker for a critical clinical functionality - namely future cognitive decline specifically within the A+SCD group. Plasma p181 tau might thus enable the identification of individuals at a higher risk of future cognitive decline already at this very early stage of the pre-dementia AD continuum.

### Plasma p181 predicts future clinical stage transition in A+SCD

The overall SCD group showed a conversion rate to MCI of 21,8% (17,0% for A-SCD and 29,5% for A+SCD) in a mean follow-up time of 3 years. The finding of 20% SCD-to-MCI converters corroborates and extends previous longitudinal studies [8, 11], suggesting that SCD can herald progressive worsening of cognitive functions for a considerable share, but not *all* SCD subjects. Numerous health issues beyond neurodegenerative diseases - including mental health conditions such as depression, anxiety, temporal stress, or even fatigue - have the potential to cause SCD. This underscores the necessity of substantiating SCD through, as shown here, - ideally peripheral (i.e. blood-based) capturable-molecular biomarkers that capture potential underlying amyloid pathology and/or neurodegeneration.

The risk of conversion to MCI was particularly increased in the A+SCD group (29,5% converters). This finding substantiates the clinical relevance of A+SCD as risk stage for further disease progression along the pre-dementia AD continuum. The average time to MCI in converters was only 2.3 years, underscoring the clinical relevance of A+SCD for designing trials for early intervention.

This higher risk of conversion to MCI was associated with higher baseline levels of plasma p181 in A+SCD, i.e. plasma p181 levels allow to predict future clinical decline and conversion to the MCI stage. This predictive association was also observed for progression to dementia in the A+MCI group. This indicates the use of plasma p181 in A+SCD as a stratification biomarker for identifying which SCD subjects might be at higher risk of converting to MCI. In addition, it suggests that plasma p181 tau – with treatment-responsivity now increasingly demonstrated [4, 37]-might also serve a as a potential therapy response biomarker for future treatment trials already targeting the SCD stage charged with clinical meaningfulness – as highly requested for biomarkers by the FDA [5] - as it reflects conversion to a substantially different stage in the clinical dementia continuum.

### Plasma p181 for identification of fast decliners in A+ SCD to facilitate trials with disease-modifying treatments

The cognitive decline in A+SCD is substantially less severe than in A+MCI [13, 38]. This presents a challenge for conducting trials with disease-modifying treatments in SCD, as there is often no relevant cognitive decline in the placebo-treated group. This highlights the urgent need to identify biomarkers that allow stratification of decliners within the A+ SCD group, where such trials would be feasible. Here, we demonstrate that plasma p181 could enable the blood-based stratification of a subgroup within A+SCD characterized by faster cognitive decline. This approach could specifically enrich and enhance the effectiveness of disease-modifying treatment trials within this pre-dementia AD window of opportunity.

### Limitations

Our study has several limitations. First, our biomarker results on the prediction of cognitive decline are based on interferences on a group-level, which restricts their use in memory clinics to predict cognitive trajectories at an individual level. Upon prospectively acquiring more data from individuals who have progressed from a cognitively unimpaired state through the successive stages of the clinical Alzheimer’s disease continuum in the ongoing DELCODE study, future analyses are warranted to allow focussing on individual longitudinal biomarker trajectories and assessing the relevance of both intra- and inter-individual variances in robustly forecasting cognitive traits. Secondly, the representativeness for the general population is limited because the individuals were recruited from specialized memory clinics where they sought help because of memory complaints. However, this characteristic is linked to a higher likelihood of including participants who will undergo objective cognitive decline in SCD, compared to those with SCD who do not seek medical assistance [8, 39]. Moreover, we consider this group to be particularly motivated to participate in research and intervention, thus representing an ideal target population for clinical trials and early treatment. The lack of significance of MRI findings in our study may stem from the constrained observational follow-up period - while longitudinal capture of 4 years follow-up biomarker and MRI volumetric data already covers a relevant timeframe, it might be too short to identify robust associations. While acknowledging this constraint, we recognize the potential for further investigation into the predictive abilities of brain atrophy in relation to blood-captured molecular pathology, particularly as extended observational follow-up times become accessible. Additionally, measurements of other plasma phospho tau species, notably p217, were not available during the analysis of this study. However, they will be assessed in future studies within this cohort.

## Conclusion

In summary, our research offers strong support for the significance of plasma p181 levels as a molecular marker of AD disease progression within A+SCD, identifying it as a distinct pre-dementia stage of Alzheimer’s disease (stage 2). This stage falls between the asymptomatic stage 1 (A+CU) and the prodromal stage 3 (A+MCI) within the NIA-AA clinical AD continuum framework. The predictive capability of plasma p181 for future cognitive deterioration and the transition to MCI at such an early phase of the disease, where not only cognitive functions are still largely intact, but also axonal degeneration is yet still just incipient (as indicated by our NfL findings) offers significant potential for the early detection and timely intervention. This includes the identification of fast decliners within the A+SCD group, making it possible to conduct more effective disease-modifying trials.

## Author Contributions

MS, DM and FJ conceived the project, designed, and supervised the research. MS and DM wrote the initial draft of the manuscript. JMO and ES assisted with analysis of samples. AL and MW measured plasma p181 and NfL. DB assisted with analysis of the imaging data. DELCODE researchers conducted clinical assessments of all subjects who participated in this study, and curated CSF biomarker and clinical data. All authors contributed to writing the manuscript. FJ is the principal investigator and ED is the co-principle investigator of DELCODE.

## Data Availability

All data produced in the present study are available upon reasonable request to the authors

## Acknowledgments

We are grateful to all participants and their families.

## Declaration of Generative AI and AI-assisted technologies in the writing process

During the preparation of this work the authors used OpenAI (2024) ChatGPT (Version 4.0) in order to improve readability and language. After using this tool/service, the authors reviewed and edited the content as needed and take full responsibility for the content of the publication.

## Supplement

### Supplemental results and discussion

#### CSF p181 mirrors plasma p181 in A+SCD vs A-SCD, but not in A+CU vs A+SCD

The cross-sectional biomarker findings for CSF p181 were similar to those for plasma p181 (Fig. S1C); however, CSF p181 could not distinguish between the A+CU and A+SCD groups. Conversely, it showed an increase in the A+MCI group compared to the A+SCD group, where plasma p181 levels had already flattened out. This suggests that plasma p181 may be more sensitive in detecting p-tau pathology in A+SCD. However, further studies are needed to replicate these results.

#### Baseline levels of plasma p181 are associated with hippocampal volume loss in A+MCI, but not in A+SCD

Hippocampal volumes at baseline were lower in SCD compared CU (3061±21 vs 3148±28 mm³, p=0.016), and even further reduced in MCI individuals (2885±36 mm³, p<0.001, Fig. S2). Within A+MCI, but not yet in the A+SCD, baseline ptau181 levels showed a qualitative association with lower hippocampal volume (A+MCI: p=0.119, A+SCD: p=0.439). Baseline NfL levels were not associated with lower hippocampal volume in any of the pre-dementia stages (Fig. S3).

## Supplemental figure captions

**Supplemental Figure 1.** The CSF Aβ42/40 ratio separates A+ from A-clinical groups, and CSF p181 replicates findings in plasma p181 in A+SCD vs A-SCD, but not between A+CU vs A+SCD. Baseline CSF of CU (blue), SCD (red), MCI (green), and AD (purple) was analyzed using ELISAs to measure (A) the Aβ42/40 ratio (B) mid-region tau (=total tau) and (C) p181. In both graphs, each point represents an individual subject, and means ± SEM are indicated. Differences between groups were assessed using Welch-ANOVA followed by Dunnetts post hoc test, \**p*<0.05, \*\**p*<0.01, \*\*\**p*<.001, \*\*\*\**p*<0.0001, n.s., non-significant. (A) The CSF Aβ42/40 ratio, (B) CSF mid-region tau, and (C) CSF p181 separates A-SCD from A+SCD and is increased in A+MCI compared to A+SCD, but not in A+SCD vs A+CU.

**Supplemental Figure 2.** P**l**asma NfL levels are not associated with future cognitive decline in A+SCD. **(C)** Baseline plasma NfL levels predict future cognitive decline only in the MCI stage, lacking notable relevance in both A+CU **(A)** and A+SCD **(B)**. Trajectories are modeled as outlined in Figure 3, incorporating an interaction term of time and baseline NfL levels.

**Supplemental Figure 3.** H**i**ppocampal volume loss in A+MCI is associated with higher baseline levels of plasma p181. Longitudinal trajectories of whole hippocampal volumes of **(A)** CU (blue), **(B)** SCD (red), and **(C)** MCI (green) individuals, stratified by CSF amyloid-positivity are depicted. Dashed black lines indicate the trajectories for the clinical groups irrespective of the CSF amyloid status. Reduced hippocampal volumes are noted in SCD compared to CU individuals. Subsequently, hippocampal volumes experience further reduction in the MCI stage, reaching their lowest levels in A+MCI. In panel D – F, baseline (BL) plasma p181 levels of the A+CU, A+SCD, and A+MCI group, categorized into a low (light grey), mid (medium grey), and high (dark grey) tertile, were used to predict whole hippocampal volumes at baseline and longitudinally over the study period of four years. In the A+MCI group, higher baseline ptau181 levels are qualitatively associated with reduced hippocampal volume. Trajectories are derived from linear mixed models with an interaction term for time and baseline p181 levels, and adjusted for age at baseline, sex, and total brain volume.

**Supplemental Figure 4.** Hippocampal volume loss is not correlated with baseline NfL. **(A-C)** Higher NfL levels at baseline are not associated with lower hippocampal volume in pre-dementia AD stages. Trajectories are modeled as outlined in Figure 4, but with an interaction term of time and tertiles of baseline NfL levels.

**Supplemental Figure 5.** Higher NfL levels at baseline do not predict clinical stage transition in A+SCD and A+MCI. Survival curves for (A) A+SCD and (B) A+MCI indicate, that higher NfL levels to not predict future progression to (A) MCI or (B) dementia. Statistical interference and labeling of the graphs are as outlined in Figure 5.

## References

1. Jia, J., et al., Biomarker Changes during 20 Years Preceding Alzheimer’s Disease.N Engl J Med, 2024. 390(8): p. 712–722.

2. Leuzy, A., et al., Blood-based biomarkers for Alzheimer’s disease.EMBO Mol Med, 2022. 14(1): p. e14408.

3. Hampel, H., et al., Blood-based biomarkers for Alzheimer’s disease: Current state and future use in a transformed global healthcare landscape.Neuron, 2023. 111(18): p. 2781–2799.

4. Hansson, O., et al., Blood biomarkers for Alzheimer’s disease in clinical practice and trials.Nat Aging, 2023. 3(5): p. 506–519.

5. FDA, Early Alzheimer’s Disease: Developing Drugs for Treatment Guidance for Industry. https://www.fda.gov/regulatory-information/search-fda-guidance-documents/early-alzheimers-disease-developing-drugs-treatment, 2024.

6. Jack, C.R., Jr., et al., NIA-AA Research Framework: Toward a biological definition of Alzheimer’s disease. Alzheimers Dement, 2018. 14(4): p. 535–562.

7. Janssen, O., et al., Characteristics of subjective cognitive decline associated with amyloid positivity. Alzheimers Dement, 2022. 18(10): p. 1832–1845.

8. Jessen, F., et al., The characterisation of subjective cognitive decline.Lancet Neurol, 2020. 19(3): p. 271–278.

9. Verlinden, V.J.A., et al., Trajectories of decline in cognition and daily functioning in preclinical dementia. Alzheimers Dement, 2016. 12(2): p. 144–153.

10. Kryscio, R.J., et al., Self-reported memory complaints: implications from a longitudinal cohort with autopsies. Neurology, 2014. 83(15): p. 1359–65.

11. Mitchell, A.J., et al., Risk of dementia and mild cognitive impairment in older people with subjective memory complaints: meta-analysis. Acta Psychiatr Scand, 2014. 130(6): p. 439–51.

12. Slot, R.E.R., et al., Subjective cognitive decline and rates of incident Alzheimer’s disease and non-Alzheimer’s disease dementia. Alzheimers Dement, 2019. 15(3): p. 465–476.

13. Rostamzadeh, A., et al., Progression of Subjective Cognitive Decline to MCI or Dementia in Relation to Biomarkers for Alzheimer Disease: A Meta-analysis.Neurology, 2022. 99(17): p. e1866–e1874.

14. Jessen, F., et al., Subjective cognitive decline and stage 2 of Alzheimer disease in patients from memory centers. Alzheimers Dement, 2023. 19(2): p. 487–497.

15. van Dyck, C.H., et al., Lecanemab in Early Alzheimer’s Disease.N Engl J Med, 2023. 388(1): p. 9–21.

16. Sims, J.R., et al., Donanemab in Early Symptomatic Alzheimer Disease: The TRAILBLAZER-ALZ 2 Randomized Clinical Trial. JAMA, 2023. 330(6): p. 512–527.

17. Teunissen, C.E., et al., Blood-based biomarkers for Alzheimer’s disease: towards clinical implementation. Lancet Neurol, 2022. 21(1): p. 66–77.

18. Karikari, T.K., et al., Blood phosphorylated tau 181 as a biomarker for Alzheimer’s disease: a diagnostic performance and prediction modelling study using data from four prospective cohorts.Lancet Neurol, 2020. 19(5): p. 422–433.

19. Janelidze, S., et al., Plasma P-tau181 in Alzheimer’s disease: relationship to other biomarkers, differential diagnosis, neuropathology and longitudinal progression to Alzheimer’s dementia.Nat Med, 2020. 26(3): p. 379–386.

20. Mengel, D., et al., Plasma NT1 Tau is a Specific and Early Marker of Alzheimer’s Disease.Ann Neurol, 2020. 88(5): p. 878–892.

21. Jung, Y. and J.S. Damoiseaux, The potential of blood neurofilament light as a marker of neurodegeneration for Alzheimer’s disease.Brain, 2023.

22. Preische, O., et al., Serum neurofilament dynamics predicts neurodegeneration and clinical progression in presymptomatic Alzheimer’s disease. Nat Med, 2019. 25(2): p. 277–283.

23. Mielke, M.M., et al., Plasma and CSF neurofilament light: Relation to longitudinal neuroimaging and cognitive measures. Neurology, 2019. 93(3): p. e252–e260.

24. Jessen, F., et al., Design and first baseline data of the DZNE multicenter observational study on predementia Alzheimer’s disease (DELCODE).Alzheimers Res Ther, 2018. 10(1): p. 15.

25. Albert, M.S., et al., The diagnosis of mild cognitive impairment due to Alzheimer’s disease: recommendations from the National Institute on Aging-Alzheimer’s Association workgroups on diagnostic guidelines for Alzheimer’s disease.Alzheimers Dement, 2011. 7(3): p. 270–9.

26. McKhann, G.M., et al., The diagnosis of dementia due to Alzheimer’s disease: recommendations from the National Institute on Aging-Alzheimer’s Association workgroups on diagnostic guidelines for Alzheimer’s disease. Alzheimers Dement, 2011. 7(3): p. 263–9.

27. Wolfsgruber, S., et al., Minor neuropsychological deficits in patients with subjective cognitive decline. Neurology, 2020. 95(9): p. e1134–e1143.

28. Mattsson-Carlgren, N., et al., Prediction of Longitudinal Cognitive Decline in Preclinical Alzheimer Disease Using Plasma Biomarkers. JAMA Neurol, 2023. 80(4): p. 360–369.

29. Papp, K.V., et al., Optimizing the preclinical Alzheimer’s cognitive composite with semantic processing: The PACC5. Alzheimers Dement (N Y), 2017. 3(4): p. 668–677.

30. Stark, M., et al., Relevance of Minor Neuropsychological Deficits in Patients With Subjective Cognitive Decline. Neurology, 2023. 101(21): p. e2185–e2196.

31. Benjamini, Y., Y. Hochberg, Controlling the false discovery rate: A practical and powerful approach to multiple testing. Journal of the Royal Statistical Society, 1995. 57: p. 289–300.

32. Ashton, N.J., et al., Differential roles of Abeta42/40, p-tau231 and p-tau217 for Alzheimer’s trial selection and disease monitoring. Nat Med, 2022. 28(12): p. 2555–2562.

33. Rogers, M.B. Revised Again: Alzheimer’s Diagnostic Criteria Get Another Makeover. 2023 [cited 2024 07/09/2024]; Available from: https://www.alzforum.org/news/conference-coverage/revised-again-alzheimers-diagnostic-criteria-get-another-makeover.

34. Moscoso, A., et al., Time course of phosphorylated-tau181 in blood across the Alzheimer’s disease spectrum. Brain, 2021. 144(1): p. 325–339.

35. Karikari, T.K., et al., Diagnostic performance and prediction of clinical progression of plasma phospho-tau181 in the Alzheimer’s Disease Neuroimaging Initiative.Mol Psychiatry, 2021. 26(2): p. 429–442.

36. Ebenau, J.L., et al., ATN classification and clinical progression in subjective cognitive decline: The SCIENCe project. Neurology, 2020. 95(1): p. e46–e58.

37. Budd Haeberlein, S., et al., Two Randomized Phase 3 Studies of Aducanumab in Early Alzheimer’s Disease. J Prev Alzheimers Dis, 2022. 9(2): p. 197–210.

38. Parnetti, L., et al., Prevalence and risk of progression of preclinical Alzheimer’s disease stages: a systematic review and meta-analysis.Alzheimers Res Ther, 2019. 11(1): p. 7.

39. Snitz, B.E., et al., Risk of progression from subjective cognitive decline to mild cognitive impairment: The role of study setting. Alzheimers Dement, 2018. 14(6): p. 734–742.

