## Supplementary figures and images for "Blood biomarkers confirm subjective cognitive decline (SCD) as a distinct molecular and clinical stage within the NIA-AA framework of Alzheimer’s disease"

### Fig. S1

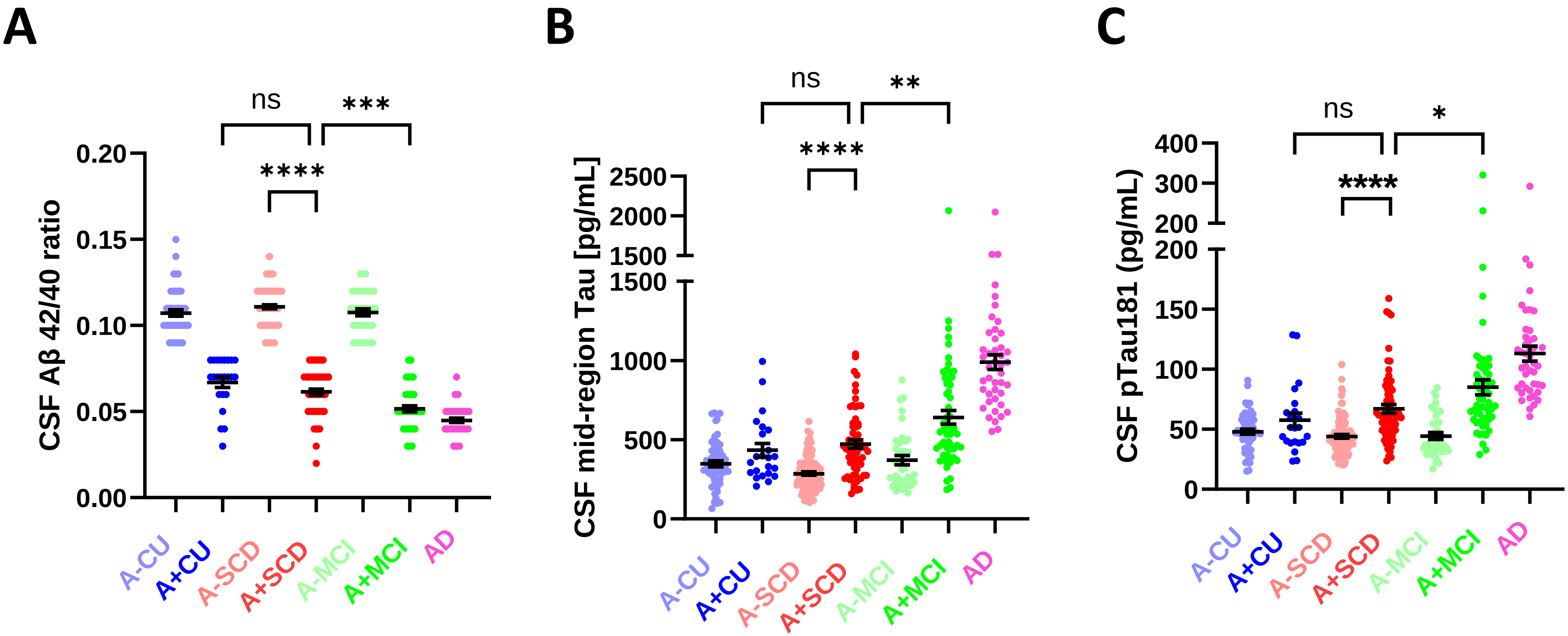

### Fig. S2

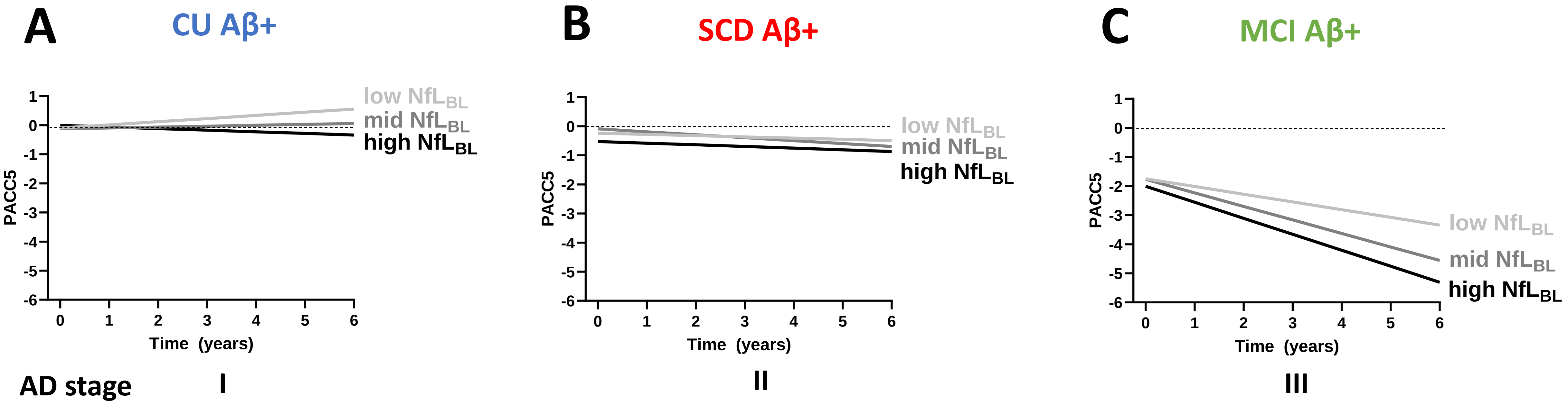

### Fig. S3

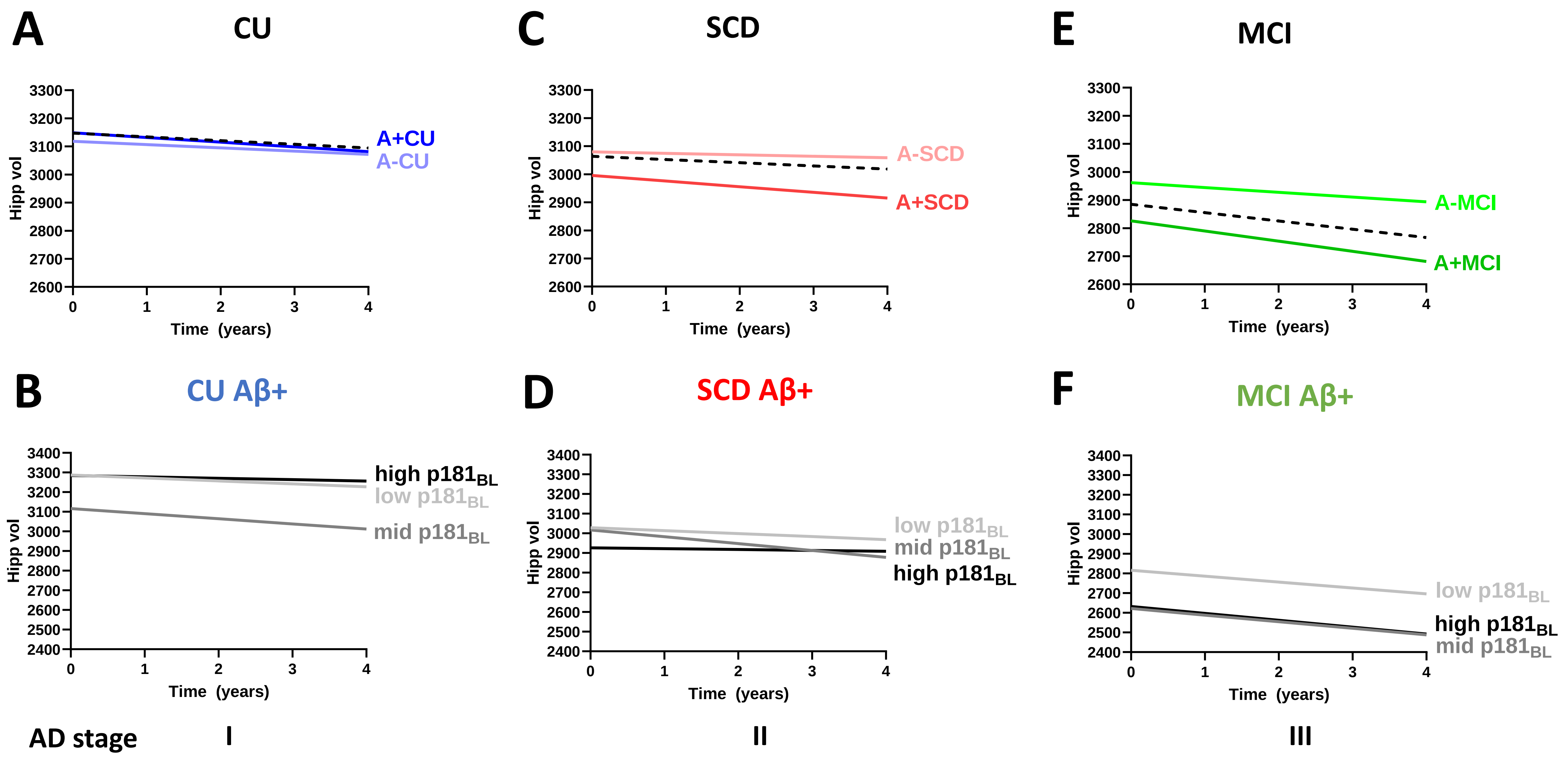

### Fig. S4

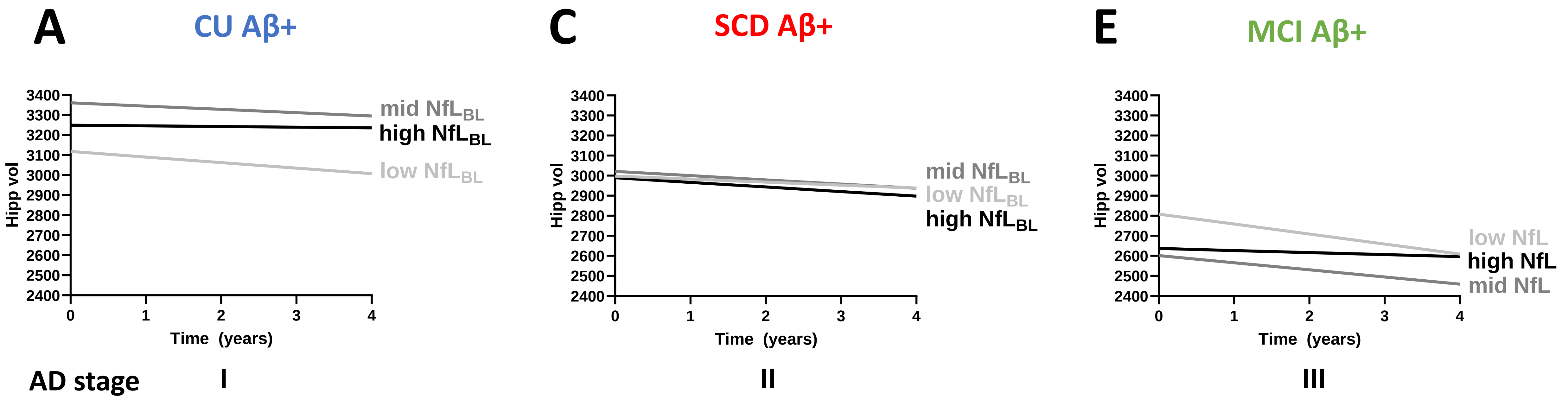

### Fig. S5

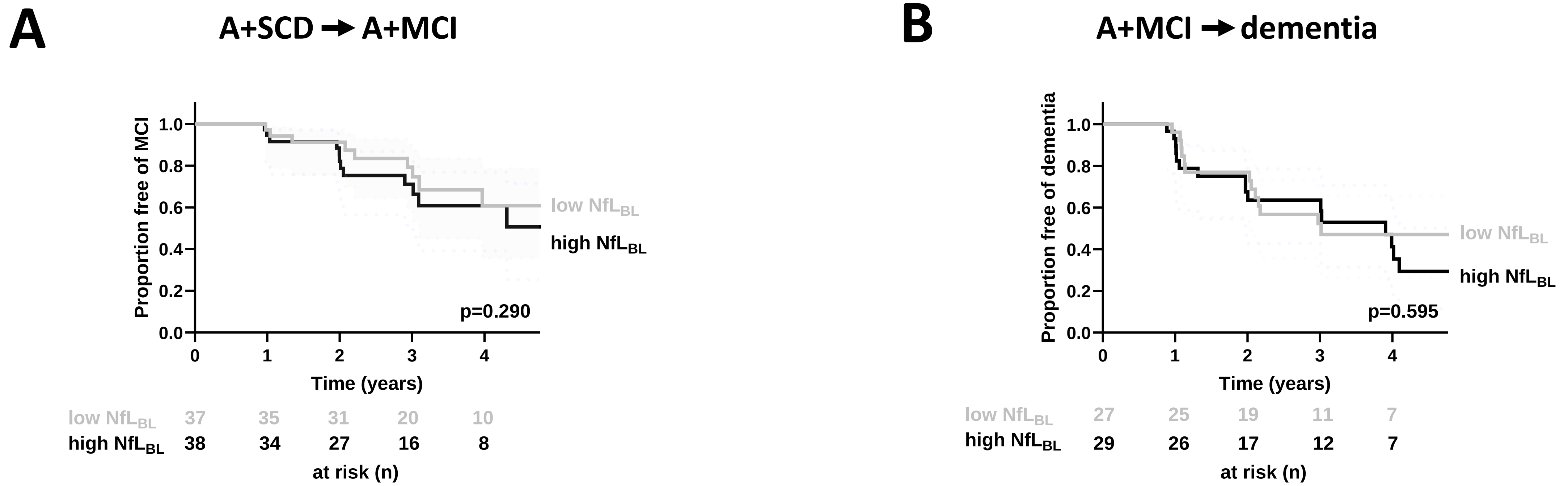
